# Cross-Cultural Adaptation and Validation of the 5C Scale to Identify Factors Associated With COVID-19 and Influenza Vaccine Hesitancy Among Healthcare Workers in Cape Town, South Africa – A Protocol

**DOI:** 10.1101/2022.06.06.22276038

**Authors:** Samuel M Alobwede, Patrick DMC Katoto, Sara Cooper, Evelyn N Lumngwena, Elvis B Kidzeru, Rene Goliath, Amanda Jackson, Charles S Wiysonge, Muki S Shey

**Affiliations:** Department of Medicine, Faculty of Health Sciences and Groote Schuur Hospital, University of Cape Town, Cape Town, South Africa; Department of Research and Innovation, Clinical division, Partners in Sexual Health, Cape Town, South Africa; Cochrane South Africa, South African Medical Research Council, Francie van Zijl Drive, Parow Valley, Cape Town, South Africa; Centre for General Medicine and Global Health, Department of Medicine, University of Cape Town, Cape Town, South Africa; Department of Global Health, Stellenbosch University, Cape Town, South Africa; School of Public Health and Family Medicine, University of Cape Town, Cape Town, South Africa; School of Clinical Medicine, Faculty of Health Sciences, University of Witwatersrand, Johannesburg, South Africa; Centre for the Study of Emerging and Re-emerging Infections (CREMER), Institute of Medical Research and Medicinal Plant Studies (IMPM), Ministry of Scientific Research and Innovation, Yaoundé, Cameroon; Centre for Research on Health and Priority Pathologies (CRHPP), Institute of Medical Research and Medicinal Plant Studies (IMPM), Ministry of Scientific Research and Innovation, Yaoundé, Cameroon; Hair and Skin Research Laboratory, Division of Dermatology, Department of Medicine, Faculty of Health Sciences and Groote Schuur Hospital, University of Cape Town, Cape Town, South Africa; Division of Radiation Oncology, Department of Radiation Medicine, Faculty of Health Science, University of Cape Town, and Groote Schuur Hospital, Cape Town, South Africa; Wellcome Centre for Infectious Disease Research in Africa (CIDRI-Africa), Faculty of Health Sciences, University of Cape Town, Cape Town, South Africa; HIV and other Infectious Diseases Research Unit, South African Medical Research Council, Durban, South Africa; Institute of Infectious Disease and Molecular Medicine (IDM), Faculty of Health Sciences, University of Cape Town, Cape Town, South Africa

**Keywords:** COVID-19 vaccines, Influenza vaccines, vaccine attitudes, vaccine confidence, vaccine hesitancy, healthcare workers, Cape Town

## Abstract

**Background:** Healthcare workers are at an increased risk of acquiring vaccine-preventable diseases and are known to be reliable source of information for the patients and their relatives. Knowledge and attitudes of Healthcare workers about vaccines are thus important determinants of their own vaccination uptake and their intention to recommend vaccinations to their patients. However, culturally adapted tools and studies to address vaccine uptake and hesitancy as well as related behaviours among Healthcare workers in the Global South are limited.

**Methods:** We propose a mixed methods project to understand the extent and determinants of vaccination hesitancy among Healthcare workers and construct a validated scale to measure this complex and context-specific phenomenon in Cape Town. We will summarise responses as counts and percentages for categorical variables and means with standard deviations (or median with inter quartile ranges) for continuous variables. We will run the Shapiro-Wilks test to assess the normality. Analysis of the variance, chi-square tests, and equivalents will be conducted as appropriate for group comparisons. Logistic regression models will also be performed to assess association between variables.

We will focus on the seasonal influenza and COVID-19 vaccine. We will use an existing tool developed and validated in Germany and the United States of America to measure five psychological determinants of vaccination (referred to as the 5C scale), as the basis to develop and validate a scale to measure the scope and determinants of vaccine hesitancy and acceptance among Healthcare workers in Cape Town.

**Discussion and conclusion:** Through this study, we hope to expand the scientific evidence based on vaccination acceptance and demand among Healthcare workers in South Africa and build resources to enable better understanding of, detection, and response to vaccination hesitancy in Cape Town.

## Background

The World Health Organization (WHO) identified vaccine hesitancy (also referred to as vaccination hesitancy) as one of the top ten threats to global health in 2019 (1, 2, 3). Vaccination hesitancy refers to the delay in acceptance or refusal of vaccines, despite availability of vaccination services. Vaccination hesitancy is complex and context-specific, varying across time, place, and vaccines (4). We use the terms vaccine hesitancy and vaccination hesitancy interchangeably in this proposal, to refer to the same phenomenon. The National Advisory Group on Immunization (NAGI) in South Africa has identified both vaccination of healthcare workers (HCWs) and vaccination hesitancy as important issues that need urgent action. In line with this plight, NAGI has formed working groups to find lasting solutions to these issues. There is thus an urgent need for local data on vaccine hesitancy and uptake among HCWs in Cape Town and South Africa as a whole.

Prior to COVID-19 pandemic, work related to vaccine hesitancy among HCWs evaluated vaccines most requested by healthcare establishments for staff members, which include influenza vaccines and hepatitis B.

HCWs were more likely to recommend vaccination if they were themselves vaccinated. A study in the United Kingdom (UK) showed that nurses who were vaccinated were more likely to recommend vaccination to their peers and other patients (17). A Nigerian study similarly reported that female nurses were more likely to recommend HPV vaccination when they expressed a willingness to be vaccinated themselves (18). Further research in Israel found that physicians were more likely to recommend vaccinations to patients if they accepted the influenza vaccine for themselves (19). Moreover, Canadian study also found that midwives who reported being immunized themselves were more likely to have trust in the safety and efficacy of influenza vaccine, and subsequently recommended vaccination to their colleagues and patients (20). It was also found tin a US study that recommending the influenza vaccine to co-workers, patients, or patients’ families, was also associated with HCWs themselves being vaccinated (21). HCWs in Iran who intended to be vaccinated against influenza in the next season were also 4.6 times more likely to recommend vaccination to their patients than those who did not intend to be vaccinated (22). However, little is known about the drivers of vaccination hesitancy among HCWs in Cape Town, and the extent of its impact on vaccination coverage; as most research on vaccination hesitancy have been conducted in high-income countries (HICs) (9, 10). In addition, tools to measure determinants of vaccination hesitancy and uptake are scarce and none of the current existing ones have been validated in South Africa. Furthermore, there are no studies on Influenza vaccine hesitancy among HCWs in Cape Town and South Africa in general. A measurement tool for vaccination hesitancy developed in Germany (known as the 5C scale, Appendix 1) has been shown to be valid and easy to administer in Europe and North America (5). The 5C scale contains a series of items (or questions) to assess five determinants of vaccine hesitancy: confidence, complacency, constraints (convenience), rational calculation, and collective responsibility (5). However, as with other vaccination hesitancy measurement tools (6, 7, 8) this scale is based on research conducted in high-income countries and has not been validated in South Africa. This potentially limits its generalizability to healthcare worker vaccination in Cape Town; hence there is need to adapt, pilot and validate this tool in our setting to determine factors associated with both Influenza and COVID-19 vaccines hesitancy among HCWs in the Western Cape, South Africa. This study aims to fill these gaps through developing capacity for vaccination hesitancy research in Cape Town and a validated measurement tool for vaccination hesitancy in this setting. The validated tool would be useful to better understand the burden and causes of vaccination hesitancy among HCWs in South Africa, monitor changes and trends over time and place in the country, and enhance the comparability of research results across the country.

## Materials and methods

### Aims, design, population, and study setting

The methodological framework for this study is adapted from Betsch and colleagues (5) and the WHO Guide to Tailoring Immunization Program (TIP) (55). Betsch and colleagues found in studies conducted in Germany and the USA that the determinants of vaccine hesitancy vary depending on the vaccine, the target group, and the country of study (5). We will focus this project on HCWs and on two vaccines, the seasonal influenza vaccine, and the COVID-19 vaccine. In this cross-sectional study, we aim to understand the extent and determinants of vaccination hesitancy among HCWs in Cape Town and to construct a validated tool to measure the phenomenon of vaccination hesitancy in this population. We will focus on two vaccines: an existing vaccine recommended for HCWs (the seasonal influenza vaccine) and the vaccine for COVID-19 for which HCWs are a priority target population. Cape Town has close to 32.479 health care workers according to (Western Cape annual report 2020/2021). By august 2021, the Western Cape Government had administered a cumulative total of 1 426 278 COVID-19 vaccines to healthcare workers, The study will take place within the Cape Metro of the Western Cape of South Africa. All healthcare facilities in the Cape Metro (whether providing integrated primary healthcare, level 2, or level 3 services) will be approached for participation in the study, making all healthcare facilities in the Cape Metro eligible for inclusion in the study and thus will avoid selection bias in the recruitment of participants. The City of Cape Town is home to about 4.4 million people, making it the second metro by population size in South Africa and the Metro also prides itself as the tourism hub of the country. Cape Town has a coastline of 294km and is also bounded by the Atlantic Ocean to the south and west. The city positions itself in Table Bay, bottom end of the African continent. It is the second largest city in South Africa and the capital of Western Province.

Globally, as of 22 May 2022 according to the WHO COVID-19 dashboard [66], there have been 524 339 768 confirmed cases of SARS-CoV-2 infections, the virus that causes COVID-19, including 6 281 260 deaths from COVID-19. A total of 11 752 954 673 vaccine doses have been administered globally by 22 May 2022. In South Africa by 22 May 2022, there have been 3 915 253 confirmed cases of SARS-CoV-2 infections with 100 898 deaths from COVID-19 reported to WHO. As of 22 May 022, a total of 35 744 528 vaccine doses have been administered in South Africa. In the Western Cape Province, a total of 701 607 confirmed cases of SARS-CoV-2 infections were reported with 22 065 deaths from COVID-19.

### The 5C scale tool

The five psychological antecedents of vaccination in the 5C scale are confidence, complacency, constraints or convenience, rational calculation, and collective responsibility (5). *Confidence* refers to the trust in (i) the effectiveness and safety of vaccines, (ii) the system that delivers them, including the reliability and competence of the health services and health professionals, and (iii) the motivations of policy-makers who decide on the need of vaccines (5). The authors of the 5C scale found in Europe and North America that individuals who lack confidence hold negative attitudes towards vaccinations, do not believe that vaccines are safe and effective, and do not trust the system that provides vaccination (5). *Complacency* occurs when “perceived risks of vaccine-preventable diseases are low and vaccination is not deemed a necessary preventive action”(5). Individuals scoring high in complacency do not perceive any risks from vaccine-preventable diseases. *Constraints* refer to practical barriers such as costs and lack of time or access. *Rational calculation* of risks and benefits of vaccination involves extensive searching for information about vaccination. *Collective responsibility* describes the situation whereby people understand the value of, and engage in vaccination, to contribute to herd immunity (5).

### Phases of the study

Our study will be conducted across four phases as described below

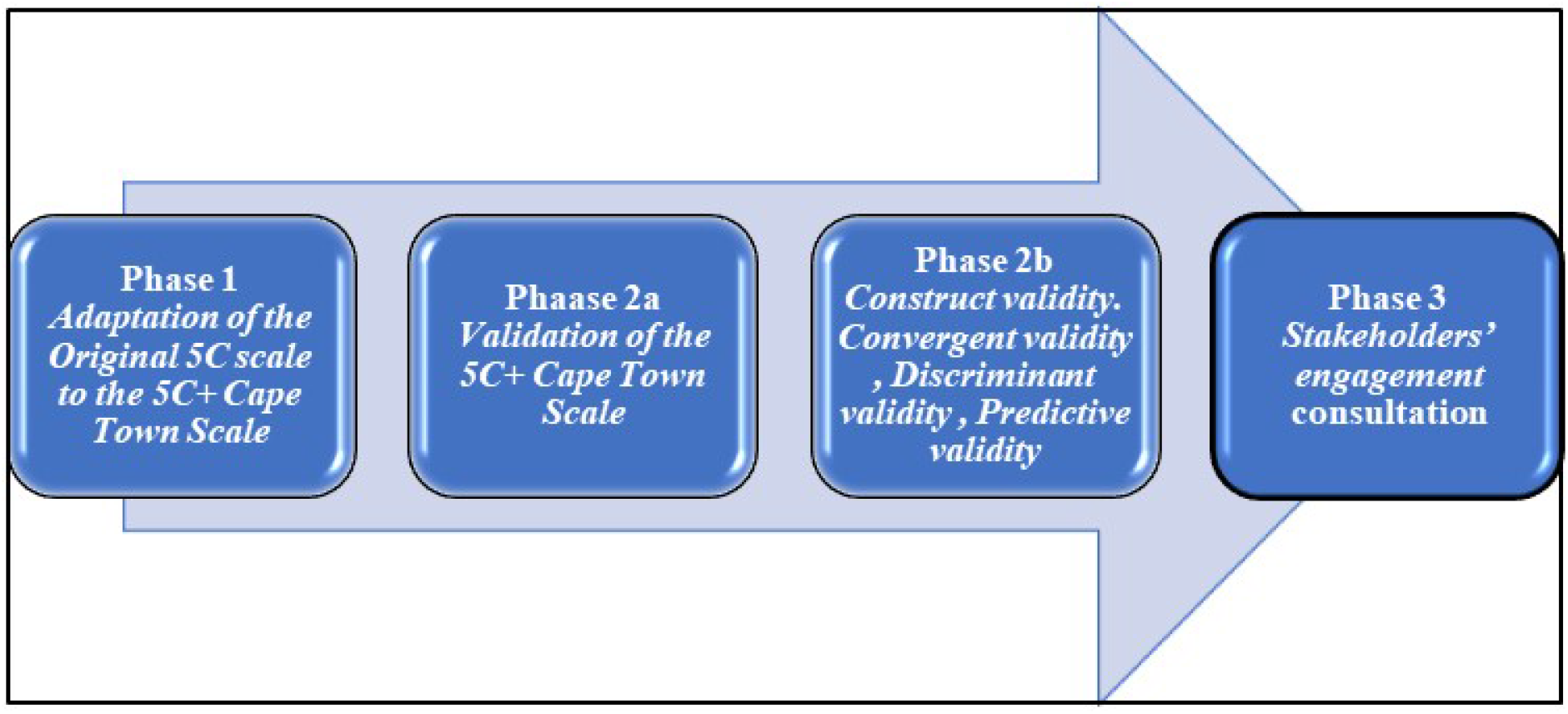

#### Phase 1: Adaptation of the Original 5C scale to the 5C+ Cape Town Scale

In this phase 1, we aim to gain deeper understanding of vaccine hesitancy among HCWs in Cape Town by engaging with relevant stakeholders. Through such engagement the study team will review the existing items in the 5C scale (6, 7, 8). This will further inform the identification and generation of additional factors and corresponding items in Cape Town, South Africa (5C+ Cape Town Scale). The expanded 5C+ Cape Town Scale will be translated into English, isiXhosa, and Afrikaans (Phase 1a). Based on our experience in the region, the added items will include willingness to be vaccinated and the compatibility of vaccination with religious beliefs. We will send the adapted 5C scale (the 5C+ Cape Town Scale) to at least five vaccine experts and HCWs in South Africa, to get feedback on whether the items are easy to understand. This engagement will help to ensure the face validity and comprehensibility of the items of the 5C+ Cape Town Scale (56). HCWs will then complete a questionnaire on their attitudes to COVID-19 and influenza vaccines, including willingness to get vaccinated (Phase 1b). The survey will be conducted both face-to-face and web-based in accordance with the COVID-19 regulation in place.

#### Phase 2: Validation of the 5C+ Cape Town Scale

The objective of this phase is to assess the reliability, construct, concurrent, and predictive validity of the adapted 5C scale for HCWs (the 5C+ Cape Town scale), and to develop a short scale (61) (Phase 2a). The validation procedure is necessary to ensure that the items assessing one of the original 5C factors produce similar results after adaptation in phase 1a and 1b as compared to the original version. Based on the findings, we may further refine the 5C items for the Cape Town specific scale (5C+ Cape Town) for further testing (Phase 2b). Construct validity queries whether the tool assesses what it aims to assess. Convergent validity will refer to the extent to which a measure correlates with other indicators of the same construct. Discriminant validity will refer to the extent to which a measure does not correlate with indicators of other constructs that are theoretically or empirically distinct. Concurrent validity will involve relating a measure to criteria assessed at the same time as the measure itself. Predictive validity will encompass associations with criteria that are assessed at some point in the future (5).

#### Phase 3: Stakeholders’ engagement

The objective of Phase 3 will be to share, discuss and obtain feedback on the findings from phases 1 and 2, including how the findings can be promoted for use within policy, practice, and research. We will identify and generate a list of key groups and individuals involved in the design, planning and implementation of healthcare worker vaccination (17). This will include policymakers, program and facility managers, professional associations, oversight, and advisory bodies, HCW unions, and HCWs themselves. The list will be developed iteratively by the research team, drawing on their collective knowledge of the relevant stakeholders, scoping of the literature, and team discussions. The methodology for developing the stakeholder list will be guided by the methods developed in a previous study to identify a complex multiple stakeholder reference sample (20)

Engagement workshops will then be conducted with a selection of individuals from identified stakeholder groups. We will employ a maximum variation purposive sampling approach (21) to ensure that workshop participant groups comprise ‘information-rich’ cases (i.e. individuals who are especially knowledgeable about or experienced in HCW vaccination) and reflect wide ranging socio-demographic characteristics and stakeholder groups.

The stakeholder engagement workshops will be conducted face-to-face or online, depending on existing Covid-19 guidelines. Each workshop will include approximately 10-15 participants. The number of workshops conducted will be determined iteratively, depending on the response rate amongst stakeholders and the findings that emerge.

The workshop will begin with a presentation of the findings from phases 1 and 2. Thereafter participants will be encouraged to provide feedback, including their views about: the extent and determinants of vaccination hesitancy among HCWs identified by the study, whether these findings resonate (or not) with their own experiences, potential issues not identified by the study, the 5C+ Cape Town Scale (including its acceptability and usability in their local settings), as well as the potential implications of the study findings. The discussions will also obtain participants views and experiences regarding how the findings can be promoted for use within policy, practice, and research to strengthen the immunisation program among HCWs. We will use multiple participatory methods to gather participant input and facilitate two-way engagement between the researchers and stakeholders (22) Potential techniques to be used include, brainstorming, meta-planning, listing, mind mapping, and ranking exercises and will comprise both large group discussions and small break-out group work. We will carefully pre-plan the composition of the small groups, considering the power-dynamics between stakeholder groups and how the views of more marginalised individuals can be properly expressed.

With the permission of participants, all workshops will be audio-recorded and transcribed verbatim, and all personal identifying information will be removed from transcripts. The anonymised transcripts will be downloaded into NVivo, a software programme that aids with the management and analysis of qualitative data. The data will be analysed through a thematic analysis, using the phases described by Braun and Clarke (23). The analysis will identify key themes regarding stakeholders’ views about the findings and strategies to enhance their uptake, as well as their views about the 5C+ Cape Town Scale and its usability. One researcher will lead the analysis, with regular discussion and feedback from other researchers.

### Healthcare workers recruitment and sample size estimation

All HCWs working in Cape Town will be eligible for inclusion. This will include nurses, medical doctors, pharmacists, hospital administrative personals, health researchers etc. Permission has been received from all the relevant authorities (including the University of Cape Town Human Research Ethics Committee, the Western Cape Provincial Department of Health. We will contact facility managers who will assist in identifying potential participants. The purpose of the study will be explained to both the managers and the potential participants.

The total sample size for the validation survey will be at least 300 HCWs, based on best practice recommendations for scale validation and performance of exploratory factor analyses (63). Given that the assessment of validity is based on the inspection of correlation patterns, studies suggest using a sample comprising at least 300 participants, which will allow detecting small correlations (r=0.2) with at least 95% power (62). While we will aim to recruit a sample size that is diverse and representative of HCWs in Cape Town, we are also mindful of the fact that representative sampling is a very complex endeavour and highly context-specific among HCWs during the pandemic period (64, 65).

### Data collection and data Management

This study will have no direct impact on clinic and hospital healthcare operations. The study will be performed with minimal direct contact with staff at healthcare facilities.

Healthcare workers will complete the consent form and questionnaire either as a hardcopy (provided to healthcare facilities) or through an online platform whereby the data will be directly captured onto the Research Electronic Data Capture (REDCap) database. The duration for completion of the consent form and questionnaire will be about 20 minutes which can be done at the convenience of the HCW. The 5C questionnaire designed to capture 23 exploratory variables including socio-demographic characteristics and attitudes of HCWs towards COVID-19 and influenza vaccination has been designed (Appendix 2).

### Expected outcome

The primary outcome of this study is a validated tool to measure vaccination hesitancy and acceptance among HCWs in Cape Town, with focus on COVID-19 and influenza vaccines. The adapted and validated long and short scales have the potential to be applicable to other parts of South Africa, to other target groups in the country, and to vaccination programs in general. This would be an important contribution to science, as no tool to measure the determinants of vaccination hesitancy and acceptance among HCWs has been previously tested in an African country. The secondary outcome will involve assessing the proportion and related risk factors of COVID-19 and influenza vaccines hesitancy and acceptance among HCWs. Lastly, the tertiary outcome will evaluate the process, barriers, and enablers of implementing the 5C+ Cape-Town scale in South Africa and other similar health settings in the global south.

### Statistical analyses

In the analyses, we will first assess the reliability of the 5C+ Cape Town scale (by assessing Cronbac’s alpha) and then determine its validity, i.e., find out whether it measures what it intends to measure. For this, we will investigate whether the 5C+ factors are associated with theoretically similar constructs (e.g. confidence should correlate with attitude) (5). To do so, we will calculate Pearson’s correlations between each 5C+ factor and the validation constructs. The pattern of results will inform the item selection process – both reliability and validity will be optimized in this process.

Then we will assess which Cape Town 5C+ factors are associated with vaccination behaviour or intention, and how strong these relations are. For vaccination behaviour we will find out for each participant if he/she has previously been vaccinated against seasonal influenza and if he/she intends to take the COVID-19 vaccine and influenza vaccine during the next influenza season. For the survey, we will categorise participants as willing or unwilling to vaccinate when a vaccine will be available to them. For descriptive analysis, we will summarise responses as counts and percentages for categorical variables and means with their standard deviations (or medians and respective inter quartile ranges) as appropriate for continuous variables. We will run the Shapiro-wilks test to assess the normality. Analysis of the variance (ANOVA), chi-square tests, and equivalents will be conducted as appropriate for group comparisons. Logistic regression models will also be performed to assess the association between each item of the 5C instrument and COVID-19 and influenza vaccines hesitancy and acceptance.

Finally, we will develop a short scale by selecting one item per subscale that best represents each subscale. The one item that correlates with the validation construct in the same direction and to a similar extent as the whole scale will be selected. As a result of this process, we will have a validated long and short scale for measuring vaccine hesitancy and acceptance among HCWs in Cape Town, each having several subscales that capture the psychological determinants of vaccination in a culturally appropriate and adapted way. Spearman’s rank correlation coefficient will be used to calculate concurrent validity between the 5C original scale and the 5C+ Cape Town long and short scales. A two-sample t-test will be used to compare the 5C+ Cape Town group scales/scores to the original. For nonparametric data, Spearman’s rho, Inter-Class Correlation Coefficient (ICC), and Wilcoxon paired test will be used for reliability assessment. Cronbach’s alpha will be used to determine internal consistency for the total score. We will consider a p-value < 0.05 to indicate statistically significant results. All analyses will be performed in R Foundation for Statistical Computing, Vienna, Austria)) V4.0.5 software.

### Ethical declaration

Ethical approval for this study was obtained from the University of Cape Town Human Research Ethics Committee (HREC Ref# (858/2020.). Furthermore, administrative permission has been obtained from the Western Cape Provincial Department of Health (WC, (# WC_202101_014), and the management of the various hospital where data will be collected. The study objectives will be clearly explained in an information sheet, before the consent form (Appendix 3) is signed and each participant will be made to know that they can discontinue their participation at any time. Data will be used only from participants who signed the consent form before participation. Confidentiality will be maintained by collecting only participant demographics without names and other participant information protected. Further to that, only age ranges will be used for further data processing with no information that my lead to identification of any participant. The risks associated with this study are minimal for participants.

## Discussion and conclusion

Our study builds on the 5C scale described previously (5). The 5C is suitable for this study because it captures multiple dimensions of vaccine hesitancy; extending the scope of existing vaccine hesitancy measuring scales that focus primarily on confidence-related aspects. Another advantage of the 5C scale is that it assesses five determinants of vaccination with only 15 questions, and a shorter version of only five questions (5). Adapting and piloting this instrument in our setting will then allow us to understand whether the 5C scale is applicable to HCWs in South Africa. This question matters because studies in Western countries have shown that the determinants of vaccine hesitancy vary depending on the target group, the country of study, and the vaccine (5) and that vaccine hesitancy among HCWs might reduce effort to curb the ongoing pandemic.

### Vaccine decision making process and HCWs

Vaccination is often cited as one of the most effective ways of controlling infectious diseases (11). However, while most people vaccinate according to the recommended schedule, this success is being challenged by individuals and groups who choose to delay or refuse vaccines (12). HCWs have had different responses to this changing environment. Certain health facilities have dropped patients if they refuse vaccination (13, 12), making the case that it puts their other patients at risk, while in other hospitals or clinics, some HCWs decided to delay vaccination or administer partial doses, in order to protect the trust relationship with their patient, although recognizing there is no clinical evidence to support such an approach (14). The vaccination decision-making process includes people who agree to be vaccinated, because they see it as the norm, and those that take their time weighing up the pros and cons of vaccination, talking with family, friends, or members of their community, searching the internet, and asking their HCWs for advice. The relevance of HCWs vaccine hesitancy is a process that has been well documented (15). HCWs are one of the strongest influences in vaccination decisions and it is on this basis that this study is grounded. Despite variations in reasons for hesitancy across geographies and vaccines among HCWs hesitancy, there are common themes that emerge globally (12, 16).

The WHO Strategic Advisory Group of Experts on Immunization (SAGE) convened a working group to investigate the nature and scope of vaccine hesitancy, which has created a model of determinants of vaccine hesitancy organized around three domains, namely: (1) contextual influences, (2) individual or social group influences, and (3) vaccine and vaccination-specific issues (12). All three domains include the influence of others on vaccine hesitancy. Domain 1 (contextual influences) includes influential leaders and individuals; domain 2 (individual or social group influences) includes personal experience with and trust in the health system and provider; and domain 3 (vaccine and vaccination-specific issues) includes the role of HCWs which is the focus of this study. In a study about HPV vaccination in Cameroon, the most important factors considered among nurses when deciding to recommend the vaccine included understanding the effectiveness and safety of the vaccine (27). In another example, a study on HCWs vaccine hesitancy in the USA found that those who were not confident in the vaccine’s efficacy and protective value were less likely to recommend it to patients or accept it themselves (21). Irrespective of the influence of HCWs vaccination behaviour on peers and others, the attitudes of HCWs towards vaccination may have a powerful influence. For example, a study about doctors’ attitudes towards measles, mumps and rubella (MMR) vaccination in Denmark illustrated that the average vaccination rate in practices with unreservedly positive attitudes about vaccination was 85%, compared with 69% in practices with more guarded attitudes (23).

### Strengths and limitations of the study

#### Study strengths

With a broad range of scenarios, it is expected that the research will generate evidence that is potentially applicable to other parts of South Africa. This study will produce relevant knowledge and has the potential for effecting policy changes (56). A complex web of influences and interactions affect the degree to which evidence is used by policymakers and managers. Among the many critical factors, the involvement of, and buy-in from, policymakers and managers at an early stage in the research is an essential element for ensuring that emerging evidence can be disseminated in timely and appropriate formats. This is the reason why we have already started engagement with the management of the healthcare facilities in Cape Town Metro.

Vaccine hesitancy is complex and context-specific, varying across time, place, and vaccines. It may be related to a specific product such as a new vaccine, new vaccine formulation, or new route of administration of a vaccine. Most of the research on the topic comes from HICs and not much is known about this issue in low and middle-income countries generally and South Africa specifically.

This study also draws on the WHO Guide to Tailoring Immunization Program (TIP) (55). The TIP framework takes a broader approach to other measurement tools and helps to 1) identify and prioritize vaccine hesitant populations and subgroups, 2) diagnose and distinguish the demand and supply–side barriers to vaccination in these populations, and 3) design evidence-informed responses to vaccine hesitancy appropriate to the setting, context, and hesitant population. This framework will help us to understand HCW vaccine hesitancy and better distinguish it from the broader range of barriers and facilitators facing HCWs vaccination.

#### Study limitations

The study limitations include that if participants lack understanding of certain questions, they will not complete the entire questionnaire for those that will complete the questionnaire online. This may lead to some exclusions for incomplete data. The second limitation is that the study will be cross-sectional assessing vaccine hesitancy at a single time point. There is possibility that attitudes of HCWs about vaccine can change over time.

### Data availability and dissemination plans

#### Dissemination plans

The results of this study will be published in scientific journals to allow researchers from sub-Saharan Africa and other countries that have not yet tested the 5C model to be able to study vaccine hesitancy in their respective settings. Moreover, the outcomes of these study will inform HCWs information sessions within our hospitals and teaching hospitals and may further inform re-enforcement of lectures on immunisation and vaccines. The outcomes will further be summarised to inform policy makers on how to better improve campaigns for COVID-19 and influenza vaccination among HCWs.

#### Data availability Statement

Main research design and tool will be made freely available to the broader academic community and public. Focus will be placed on data management and protection of participants information as well as faithful and reproducible record keeping, with an emphasis on transparency and accountability in methods utilised. Primary outcome being identification of factors associated with vaccine hesitancy and affecting vaccine uptake by healthcare workers. The results will be documented in text, plots, and images. Findings will be legible, reasonably organized, and sufficiently detailed for future use by other researchers wishing to reproduce the outcome. Research Electronic Data Capture (REDCap) computer software will be used to capture, store and generate data. All raw data in electronic format will be stored in an organised fashion and widely used and accessible formats will be employed. All stored electronic data, updated frequently, will be continuously backed up to external (including cloud-based) media. Detail results of the main research products will appear online in digital format in text, tables, plots, and images in peer-reviewed journal articles and/or conference proceedings. Records will be durable, accessible, and made safe from tampering or falsification.

## Conclusion

In this survey of vaccine hesitancy among HCWs, we will adapt, pilot, and validate the 5C scale instrument among HCWs in South Africa. By doing so, we will gain knowledge on the extent and determinants of vaccination hesitancy among HCWs in an African context. This may facilitate other researchers to study this complex and context-specific phenomenon of vaccination hesitancy in South Africa and generally in Africa. This would expand the evidence base on vaccination hesitancy in Africa and enhance the generalizability of current theoretical causal models of vaccination hesitancy. Such knowledge will enable us and other researchers in the future to design interventions to reduce vaccination hesitancy, which are evidence-based, contextually appropriate, and focused on causal mechanisms, thus improving their effectiveness, cost-efficiency, and acceptability.

## Data Availability

Deidentified research data will be made publicly available when the study is completed and published.

## Abbreviations

5C Scale: Tool to measure five determinants of vaccine hesitancy (confidence, complacency, constraints, rational calculation, and collective responsibility)
5C+ Scale: 5C scale adapted to Cape Town
CHC: Community Health Center
GVAP: Global Vaccine Action Plan
HCW: Healthcare Workers
HICs: High Income Countries
HREC: Human Research Ethics Committee
MMR: Measles, Mumps and Rubella
NAGI: National Advisory Group on Immunization
SAGE: Strategic Advisory Group of Experts on Immunization
SWOT: Strengths Weaknesses, Opportunities, and Threats
TIP: Tailoring Immunization Program
UK: United Kingdom
UCT: University of Cape Town
USA: United States of America
WP: Western Cape province
WHO: World Health Organization?

## Appendix

1. Original 5C scale
2. Questionnaire
3. Informed consent form

